# Consolidation of a Genomic Epidemiological Surveillance Network for Tuberculosis (REVIGET) in Northern and Northeastern Brazil: A Study Protocol

**DOI:** 10.1101/2025.05.29.25328602

**Authors:** Emilyn Costa Conceição, Karla Valéria Batista Lima, Cristiane Cunha Frota, Theolis Costa Barbosa Bessa, Adriana Ayden Ferreira, Abhinav Sharma, Davi Josué Marcon, Layana Rufino Ribeiro, Alex Brito Souza, Danna Karen Corrêa dos Santos, Carlos Augusto Abreu Alberio, Ricardo José de Paula Souza e Guimarães, Ismari Perini Furlaneto, Lilian Eduarda de Oliveira Freitas, Emmily Oliveira Amador, Hendor Neves Ribeiro de Jesus, Leonardo Bruno Paz Ferreira Barreto, Maria Cristina Silva Lourenço, Tulio de Oliveira, Robin Mark Warren, REVIGET Consortium

**Author notes:** Equally contributed (Emilyn Costa Conceição, Karla Valéria Batista Lima, Robin Mark Warren). Equally supervised (Tulio de Oliveira, Robin Mark Warren). Corresponding Authors: Emilyn Costa Conceição, Karla Valéria Batista Lima.

## Abstract

Globally, tuberculosis (TB) remains a top cause of death from infectious diseases, with an estimated 1.5 million deaths annually. Given its substantial social and economic burden, TB is a priority in the United Nations 2030 Agenda for Sustainable Development. The WHO’s End TB Strategy emphasises research, innovation, and the rapid implementation of new technologies such as whole genome sequencing (WGS), which are pivotal for precision health approaches and drug-resistant TB (DR-TB) surveillance. This study aims to strengthen genomic TB surveillance in the North and Northeast Brazilian regions by applying WGS to study DR-TB cases, training professionals in genomics and bioinformatics, and deploying a national surveillance platform (GEMIBRA). This is an observational, cross-sectional, prospective, quantitative and qualitative study to be conducted in the states of Pará, Amazonas, Ceará, and Bahia. A total of 500 *Mycobacterium tuberculosis* complex (MTBC) isolates from DR-TB cases will undergo WGS, representing 87% of the expected DR-TB cases, including cases of non-tuberculous mycobacteria (NTM) isolates among the DR samples. Data will be analyzed for genotype-phenotype correlations, mutation patterns, and associations with clinical and epidemiological characteristics. Capacity-building activities, including theoretical and hands-on bioinformatics training, will be carried out. The GEMIBRA platform will support data visualization, spatial distribution, and clinical decision-making. The project will generate evidence for validate the contribution of the integration of WGS into Brazil’s TB surveillance system, support precision treatment approaches, and inform public health interventions. It will also contribute to workforce development and the introduction of decentralized WGS-based diagnostics in resource-limited regions. The project adopts a translational research model and a networked, decentralised approach, facilitating the prompt integration of the knowledge generated into public health practice. Ultimately, this work will contribute to combating TB transmission by identifying transmission dynamics, emerging resistant strains, and informing the National Plan to End TB as a public health problem.

## 1. Introduction

Tuberculosis (TB) remains a curable infectious disease that continues to cause approximately 1.5 million deaths annually worldwide. Due to its profound economic and social burden, TB is prioritized in the United Nations 2030 Agenda for Sustainable Development [1] and remains a central focus for the World Health Organization (WHO) and Brazil’s Ministry of Health. The WHO’s End TB Strategy emphasizes intensified research, innovation, and rapid adoption of advanced technologies—among them, next-generation sequencing (NGS)—to improve TB control and surveillance [2].

Adding complexity to the diagnostic landscape, non-tuberculous mycobacteria (NTM) infections, that can present clinical symptoms similar to TB but require different treatment, are increasingly being recognized as a public health concern. Correct identification of the *Mycobacterium tuberculosis* complex (MTBC) (group of bacteria that causes TB) and NTM species is essential for ensuring appropriate therapy, as treatment is highly species-specific [3–5].

In this context, NGS offers several applications in infectious diseases, the most prominent being whole-genome sequencing (WGS), which provides comprehensive coverage of the bacterial genome and suitable for transmission studies or discovery of novel mutations. In addition, targeted NGS (tNGS), such as Deeplex® Myc-TB (GenoScreen), offers high coverage sequencing of key particular genes including drug resistance associated genes and particularly interesting for detecting heteroresistance and polyclonal infections and capable of being used directly in clinical samples. WGS however remains the preferred method for high-resolution genomic surveillance [6,7].

As observed during the COVID-19 pandemic, integrating genomic technologies into public health frameworks is essential. For TB, a One Health approach is increasingly relevant, acknowledging the interplay between socio-environmental factors, host-pathogen interactions, including animals, antimicrobial resistance, and technological advancements [8,9].

Despite the introduction of new anti-TB drugs—bedaquiline, delamanid, linezolid, and pretomanid—limited knowledge exists regarding their resistance mechanisms. The WHO and scientific community advocate for expanded genomic surveillance and genotype-phenotype correlation studies. However, such strategies must be adapted to resource-constrained settings, where access to technical infrastructure and trained personnel is limited [10]. In the context of NTM, further research is needed to establish a comprehensive catalogue of resistance-associated mutations to inform treatment decisions, similar to the resources already available for *Mycobacterium tuberculosis*, the human-adapted MTBC species [11,12].

Brazil remains among the countries with the highest TB burden globally, belonging to the WHO list of high-burden TB countries (HBC) [13] and a rising clinical relevance of NTM in TB-endemic regions in Brazil is observed [14–16]. Regions such as the North and Northeast of Brazil face disproportionate challenges due to economic disparities, limited laboratory capacity, and delayed diagnosis. Studies have shown that delays in detecting drug resistance are associated with worse clinical outcomes, including death. In addition, systemic inequities rooted in poverty, structural racism, and geographic disparities undermine effective TB control in these regions [17].

It is noteworthy that these regions are also underrepresented in molecular epidemiological studies on TB and face significant limitations in infrastructure to support timely genomic diagnosis and surveillance of this disease [18], challenges that are also relevant to NTM [19]. These regions are home to traditional communities, exhibit lower TB cure rates, and have limited access to innovative diagnostic technologies [20]. Therefore, there is an urgent need for research that not only advances scientific knowledge but also strengthens local health systems through capacity-building initiatives and the development of context-specific diagnostic tools [19].

## 2. Materials and Methods

### 2.1 Study Aim and Objectives

This study aims to apply WGS to determine the drug resistance profile in cases of drug-resistant TB (DR-TB) and TB-NTM coinfection identified by the national diagnostic laboratory network, and to train healthcare professionals in data analysis, interpretation, and clinical decision-making based on internationally validated methodologies. The project will also implement GEMIBRA, an interactive web-based platform developed to support Brazil’s National Health Data Network (RNDS) in modernizing TB and TB/NTM co-infection surveillance.

The study specific objectives are: 1) To implement WGS as a diagnostic tool for resistance profiling using samples submitted through the regular workflow of the Central Laboratoraies (LACENs) and other diagnostic centres; 2) To conduct genomic and epidemiological analyses of TB and NTM infections; 3) To train laboratory professionals in the North and Northeast regions in WGS, bioinformatics, and data analysis applied to Mycobacteria through theoretical-practical courses, both in-presential and virtually; 4) To deploy the GEMIBRA platform (http://www.ioc.fiocruz.br/gemibra/) for facilitating TB surveillance and research through WGS data, including lineage/genotype identification, mutation profiling, phenotypic resistance modelling, transmission patterns, and spatial distribution; 5) To engage health professionals and decision-makers in interpreting WGS results as a potential diagnostic support tool aligned with international guidelines, fostering integration with national systems; 6) To develop educational materials (booklets, books, videos, and apps) and promote health engagement activities to increase TB awareness— particularly drug-resistant TB—among the general public, students, health professionals, and TB patients in diverse educational and social spaces; 7) To evaluate the costs associated with diagnostic methods and the turnaround time within the diagnostic workflow.

### 2.2 Study Design and Setting

This is an observational, cross-sectional, prospective, epidemiological, mixed-methods (quantitative-qualitative), descriptive and analytical study. The study will be conducted over a period of 24 months (2024–2026) in selected locations in the North and Northeast of Brazil, including Ananindeua and Belém (Pará), Manaus (Amazonas), Fortaleza (Ceará) and Salvador (Bahia). The city of Rio de Janeiro (Rio de Janeiro), located in Southeast Brazil, will serve as a reference laboratory to perform the drug susceptibility testing (DST) by determining the minimum inhibitory concentrations (MICs). The study settings are highlighted in Fig 1.

**Fig 1.**
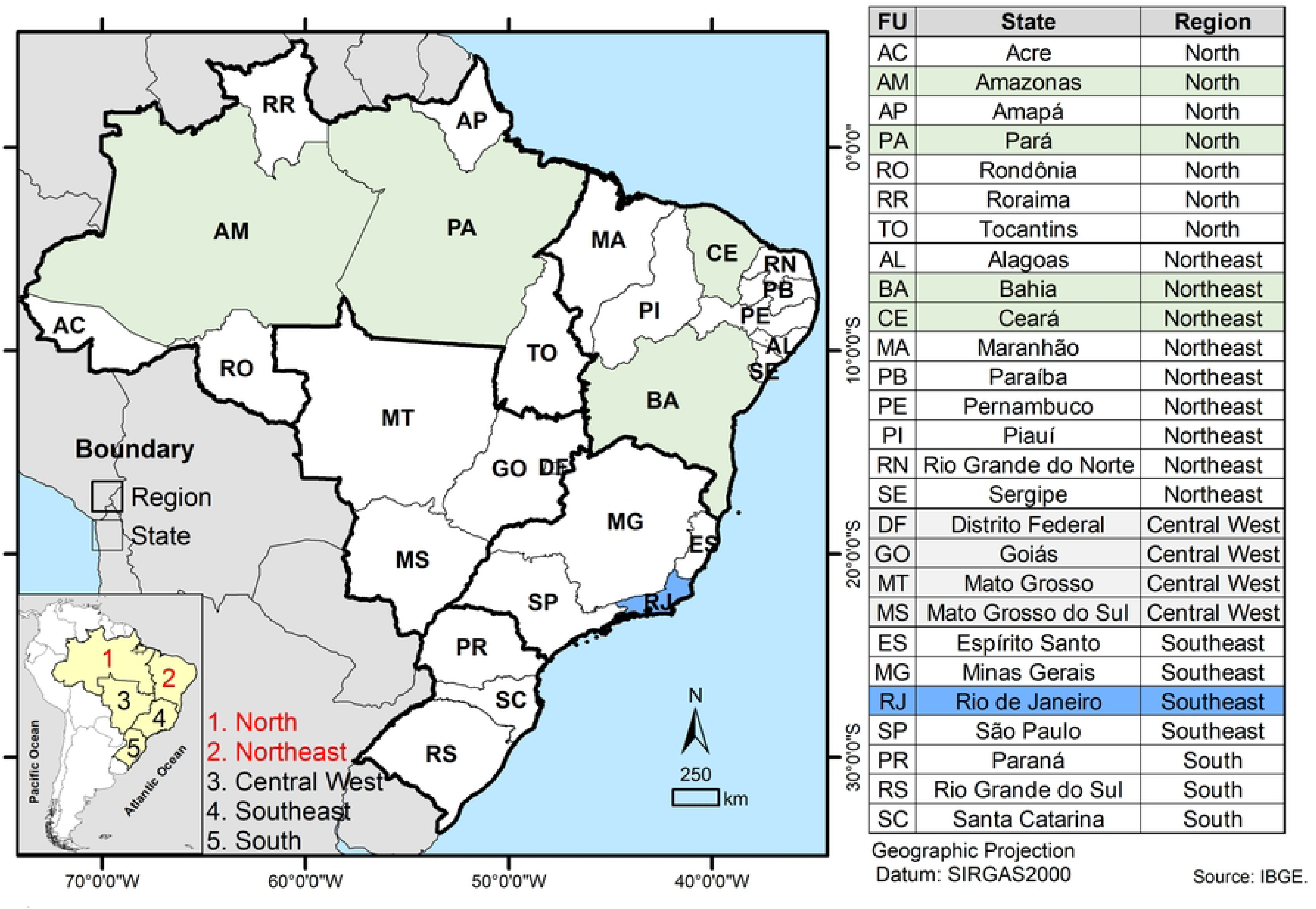
Geographic map of Brazil showing its five major regions and the 27 federal units (states). Study sites involved in sample and data collection, routine diagnostic processing, and capacity-building activities are highlighted in green as part of the North and Northeast regions. The site designated for minimum inhibitory concentration (MIC) testing is indicated in blue.

### 2.3 Sample size and Eligibility Criteria

Sampling will follow the routine diagnostic workflow and will include *M. tuberculosis* isolates from individuals diagnosed with pulmonary or extrapulmonary DR-TB, confirmed via either solid or liquid culture media. If NTM are later isolated from a sample initially identified as *M. tuberculosis*—whether by RMT or culture—the case will be further assessed to investigate potential TB/NTM coinfection or diagnostic misclassification.

Sample size estimation was based on internal data from each LACEN. In 2023, the LACENs Amazonas, Bahia, Ceará and Pará detected 176, 152, 125 and 120 DR-TB cases, respectivelly, totalling 573 DR-TB cases. Considering a coverage of a minimum 85% of DR-TB cases in 2024 as based on the data from 2023, and NTM isolated after TB detection, we propose to perform WGS of 500 samples (87%) from 2024.

Sample size estimation was informed by internal data provided by each participating LACEN. In 2023, the LACENs of Amazonas, Bahia, Ceará, and Pará reported 176, 152, 125, and 120 cases of DR-TB, respectively, totalling 573 DR-TB cases. Based on these figures and targeting a minimum coverage of 85% of DR-TB cases detected in 2024, including NTM identified following TB diagnosis, we propose conducting WGS on 500 isolates (approximately 87% of the projected total). If fewer than 500 eligible cases are identified across these sites in 2024, sample collection will be extended into 2025. Priority will be given to complex TB cases, including those with persistent treatment failure, suspected polyclonal infections, or discordant DST results.

Inclusion criteria comprise patients of any age, sex, or nationality diagnosed with any form of DR-TB, regardless of co-infections or comorbidities, provided that they are registered in one of the four study states (Amazonas, Bahia, Ceará, or Pará) and have stored samples with viable mycobacterial cultures. Exclusion criteria are being infected with drug susceptible *M. tuberculosis* or for whom no culture isolates are available.

### 2.4 Sample collection and processing

Sample collection will take place after routine diagnostic testing has been completed, between 1 January 2024 and 31 December 2025, at the time when bacterial samples are archived. If fewer than 500 DR-TB cases are enrolled across the study sites in 2024, collection will continue into 2025, with priority given to complex DR-TB cases, such as patients experiencing persistent treatment failure, suspected polyclonal infection, or discordant DST results.

The sample collection strategy takes into account the variability in routine TB diagnostic practices across different regions and states within Brazil. In general, people exhibiting clinical symptoms indicative of TB are initially referred to a primary healthcare facility, where sputum specimens are collected for diagnostic evaluation. These specimens are analyzed via microscopy, Ziehl-Neelsen (ZN) staining and, when available, the RMT GeneXpert MTB/RIF Ultra assay. Mycobacterial culture is performed on Ogawa medium at the same facility for all samples. Upon confirmation of culture positivity, isolates are forwarded to the LACEN, where each isolate is subcultured on Löwenstein-Jensen (LJ) medium. Once the culture is positive, DST is performed on the first-line drugs and samples are cryopreserved at −20°C or −70°C for traceability. When resistance to rifampicin or isoniazid is detected, then the samples are subcultured in the MGIT instrument for second-line DST and/or the RMT LPA for first and/or second-line drugs when available at the LACEN level; otherwise, they are shipped to their specific regional reference laboratory (RRL). If the test is not performed at the RRL, then it is shipped to the national reference laboratory (NRL) for further second-line DST testing.

This study will be conducted in this scenario as described in Fig 2. The WGS method will be integrated into the routine diagnostic workflow for samples showing resistance to first-line anti-TB drugs and for NTM detected in specimens initially suspected to be TB.

**Fig 2.**
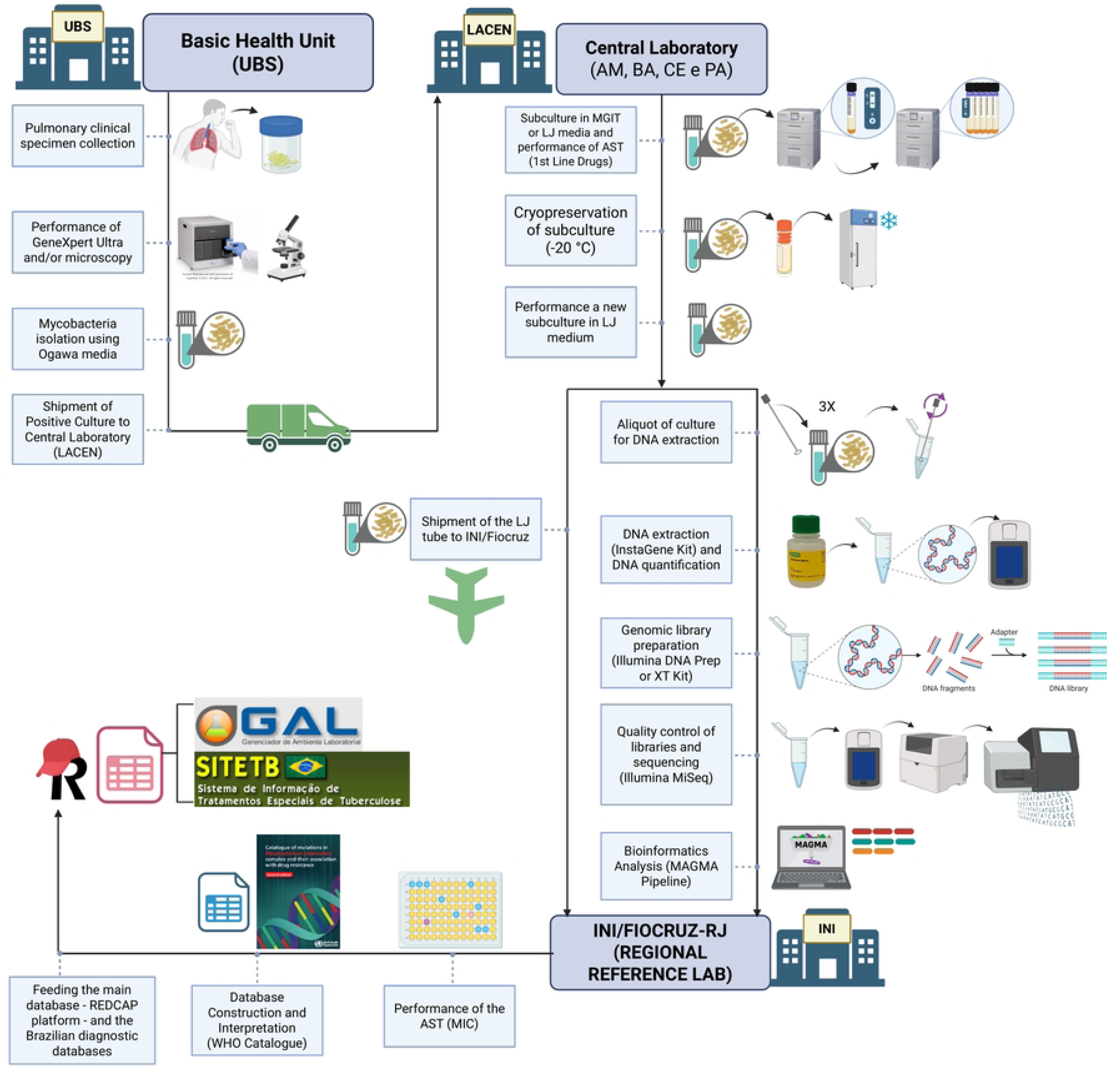
Overview of the protocol workflow from sample collection to data analysis as implemented in the Genomic Epidemiological Surveillance Network for Tuberculosis (REVIGET) study in Northern and Northeastern Brazil. DST: Drug Susceptibility Test; Basic Health Unit (UBS from the Portuguese name “*Unidade Básica de Saúde*”), LACEN: Central Laboratory from the states of Amazonas (AM), Bahia (BA), Ceará (CE) and Pará (PA); INI: Institute National of Infectiology; LJ: Löwenstein–Jensen; MIC: Minimum Inhibitory Concentration; MGIT: Mycobacteria Growth Indicator Tube. Created in BioRender. Warren, R. (2025). Created in BioRender. Warren, R. (2025) https://BioRender.com/undefined.

From the routine cryopreserved LJ culture, the Mycobacterial isolate will be subcultured on LJ tubes. An aliquot will be designated for DNA extraction and WGS, while another aliquot will be designated to perform the pDST minimum inhibitory concentration (MIC) method for the following drugs: rifampicin, isoniazid, streptomycin, ethambutol, amikacin, levofloxacin, moxifloxacin, clofazimine, bedaquiline, linezolid, delamanid and pretomanid. The MIC interpretation will follow the Clinical and Laboratory Standards Institute guideline M24 (CLSI) [21]. For the MIC test, the samples of the four sites will be shipped to the RRL at the National Institute of Infectology (INI) at Oswaldo Cruz Foundation, in a way not to interfere with the current routine system, and to centralized the phenotypic tests and maintain standardization.

The WGS will be performed locally, initially during the laboratory capacity building and later at the local site. Between 10 and 20 mycobacterial samples from each LACEN (Amazonas, Bahia, Ceará and Pará) will be used during hands-on training sessions focused on capacity building and data analysis. The remaining samples collected throughout the study period will be processed and analyzed after the training is completed.

For this, sample handling will occur in Biosafety Level 2 plus (BSL-2 +) laboratories. After heat inactivation of cultures at 80°C for 1 hour, samples will be transferred to Biosafety Level 2 (BSL-2) laboratories for downstream procedures. Laboratory personnel will strictly follow locally approved biosafety protocols and standard operating procedures, including the continuous use of personal protective equipment (PPE) and appropriate handling of reagents and specimens.

The mycobacterial DNA will be extracted from solid culture on LJ using the InstaGene (IGM) Matrix kit (Bio-Rad, USA) as per Conceicao et al. 2024 protocol with modifications, which includes the use of a Vortex for 5 minutes instead of a high-speed homogenizer instrument [22]. The DNA quantification in ng/uL will be assessed using a Qubit Fluorometer (Thermo Fisher Scientific, Waltham, USA) double-strand High Sensitive (dsHS) kit.

Following, the genomic DNA samples with a minimum of 0.5ng/uL in a volume of 150 uL IGM Matrix will be selected for library preparation using the DNA Prep kit or Nextera XT kit (Illumina, Santa Clara, USA), depending on their availability. The genomic libraries’ quality control will be assessed using a Qubit Fluorometer dsHS kit for quantification (ng/uL) and TapeStation system for fragment size in base pairs (bp) verification using a D5000 kit whose range is expected to be around 400-600 bp. The sequencing will be performed using Illumina MiSeq platforms, which are already installed at the LACENs.

The Bioinformatics analysis will use the MAGMA pipeline [23], and drug resistance interpretation will follow the WHO catalogue of mutations (Second Edition) [12]. WGS results and phenotypical profile will be interpreted using WHO’s mutation catalogue, and all validated results are subsequently uploaded to the REDCap database and Brazilian diagnostics databases Laboratory Environment Manager (GAL) (http://gal.datasus.gov.br/GALL/index.php?area=01) and Special Tuberculosis Treatment Information System (SITE-TB) (http://sitetb.saude.gov.br/).

### 2.5 Data Collection and Management Plan

Primary data will be managed by diagnostic laboratories and institutions in each participating region. Clinical, epidemiological, and sociodemographic data—anonymized via ID codes—will be obtained from the Brazilian systems GAL and SITE-TB. Data will be obtained by healthcare professionals registered with login credentials in both GAL and SITE-TB systems, and working at LACENs or reference hospitals. These professionals will select and anonymize all relevant information before sharing it with the research team for analysis. The dataset will include individuals diagnosed with DR-TB between January 2024 and December 2025. Data collection will take place from January to December 2025. All data will be stored and managed using the REDCap system (https://project-redcap.org/). Upon completion of the analysis, only anonymized genomic and phenotypic data of the bacterial strains will be made publicly available.

The project will be coordinated using the Notion platform (https://www.notion.so/) for task management and REDCap for data management. Periodic weekly remote meetings will ensure progress monitoring. Budget and administrative risks will be managed with support from the Centre of Excellence in Project Management (*Núcleo de Excelência em Gestão de Projetos*) of the Gonçalo Moniz Institute, Oswaldo Cruz Foundation, Bahia (NEGEP/IGM/Fiocruz-BA).

### 2.6 Capacity Building in WGS and Bioinformatics

Capacity building is a central component of this collaborative initiative between Brazil and South Africa. Training activities will be coordinated by two representatives from Stellenbosch University, affiliated with the TB Genomics Research Group, the Bioinformatics Unit, and the Centre for Epidemic Response and Innovation (CERI), with support from two previously trained personnel from the IEC.

Initial training sessions will be held at the state public health laboratories (LACENs) in Amazonas, Pará, Bahia, and Ceará. Each session will consist of a one-week intensive course (40 hours), featuring a public workshop conducted in Portuguese and offered both in person and online.

The training program will cover: (1) theoretical foundations of next-generation and whole genome sequencing, including their applications in public health; (2) hands-on laboratory sessions encompassing DNA extraction, library preparation, quality control, and sequencing procedures; and (3) bioinformatics modules focused on pipeline execution, data analysis, and the generation of patient-centred reports.

Follow-up training sessions will be held monthly, comprising both theoretical and practical components in bioinformatics. These will be supplemented by additional activities and assessments to reinforce learning outcomes and ensure the continuous development of trainees’ competencies and skills.

### 2.7 Development of the GEMIBRA Platform

The development of GEMIBRA (http://www.ioc.fiocruz.br/gemibra/) will occur in four stages: 1) retrieval, QC, and validation of public MTBC genomes; 2) data analysis using the MAGMA pipeline (https://github.com/TORCH-Consortium/MAGMA), which identifies lineage, genotype, resistance profile, transmission dynamics, and supports clinical decision-making; 3) integration with geospatial data to map transmission by state; and 4) development, validation, and deployment of an interactive user interface for researchers and health professionals.

### 2.8 Statistics and Data Analysis

Sociodemographic and clinical-epidemiological characteristics will be analyzed through descriptive statistics using frequencies for categorical variables, and mean and standard deviation (SD) for parametric variables with a 95% confidence interval (95% CI), or median and interquartile range (IQR) for non-parametric variables. Normality will be evaluated using Shapiro-Wilk test. A significance level of 5% (α = 0.05) will be applied. Statistical analyses will be conducted using GraphPad Prism for MAC (version 10.4.2 or later). Bioinformatics analysis will be performed using the MAGMA pipeline, which is optimized for analyzing MTBC genomes, including contaminated samples. To assess the cost of diagnostic methods, both Cost-Consequence Analysis (CCA) and Cost-Effectiveness Ratios (CER) will be applied. For the CCA, the annual acquisition cost, diagnostic accuracy, and turnaround time of each test will be compiled in a Microsoft Excel 365 matrix. The CER will be derived by comparing test costs against diagnostic quality and speed, thus informing the annual cost-efficiency of each method.

### 2.9 Ethical Considerations

This study was submitted for ethical evaluation through the Brazilian system (Plataforma Brasil) and addressed to the Institutional Review Board - Research Ethics Committee (*Comitê de Ética em Pesquisa* - CEP) of the led institution (IEC), which was approved No 7,282,078 and Certificate of Presentation of Ethical Appreciation (CAAE): 85134924.4.0000.0019. The CEP waived the requirement for informed consent due to: 1) impracticality of reaching a large number of people, as the bacterial sample and data will be collected retrospectively, and 2) requiring consent would introduce bias or compromise scientific validity to assess genetic diversity and DR-TB mutations for surveillance purposes.

Access will be granted to archived samples and associated data, including laboratory test names and results, dates of sample collection and results, clinical outcomes, and epidemiological information such as age and sex. All information will be fully anonymised before analysis. Data will be obtained from the Brazilian information systems GAL and SITE-TB, as previously described. If needed, medical records may also be reviewed. This project was also registered per Brazilian Law No. 13,123/2015 and its regulations, through the National System for the Management of Genetic Heritage and Associated Traditional Knowledge (SisGen), under Registration No. A247AD9.

### 2.10 Safety considerations

This study involves potential risks related to laboratory procedures, administrative processes, data management, and data interpretation. To ensure the safety of participants and personnel, as well as the integrity of the research, multiple preventive and mitigation strategies will be implemented.

Laboratory safety will be a priority throughout the study, and biosafety measures will be followed to ensure a safe working environment for all staff involved, following the established regulatory standard No. 32 “Occupational Safety and Health in Health Care Services” [24]. All clinical specimens and *M. tuberculosis* cultures will be processed following established biosafety regulations. The risk level in a TB laboratory refers to the likelihood of laboratory personnel becoming infected with *M. tuberculosis* due to procedures performed in the laboratory. Activities associated with a low risk involve a minimal likelihood of generating infectious aerosols from clinical specimens and typically involve a low concentration of infectious particles. Such activities include direct smear microscopy, sample preparation for automated nucleic acid amplification tests (e.g., molecular rapid test – RMT), and the swab method for bacterial isolation (culture) (Ogawa-Kudoh). These can be performed outside a Biological Safety Cabinet (BSC) class II, in case they are conducted near a Bunsen burner and with appropriate use of personal protective equipment (PPE)[25,26].

Procedures associated with a moderate risk involve a moderate potential for aerosol generation but still involve low concentrations of infectious particles. These include processing and concentration of biological samples for inoculation into primary isolation media (e.g., Löwenstein-Jensen or MGIT) and direct drug susceptibility testing, such as the Line Probe Assay (LPA), on processed sputum samples using centrifuge equipment, for example. These activities must be performed within a BSC and require proper use of PPE. High-risk activities are those with a high potential for generating infectious aerosols and involve high concentrations of infectious particles. These include manipulation of cultures grown in solid or liquid media for species identification, phenotypic drug susceptibility testing, or LPA. All such procedures must be conducted within a BSC with strict adherence to PPE protocols (Fig 3) [26].

**Fig 3.**
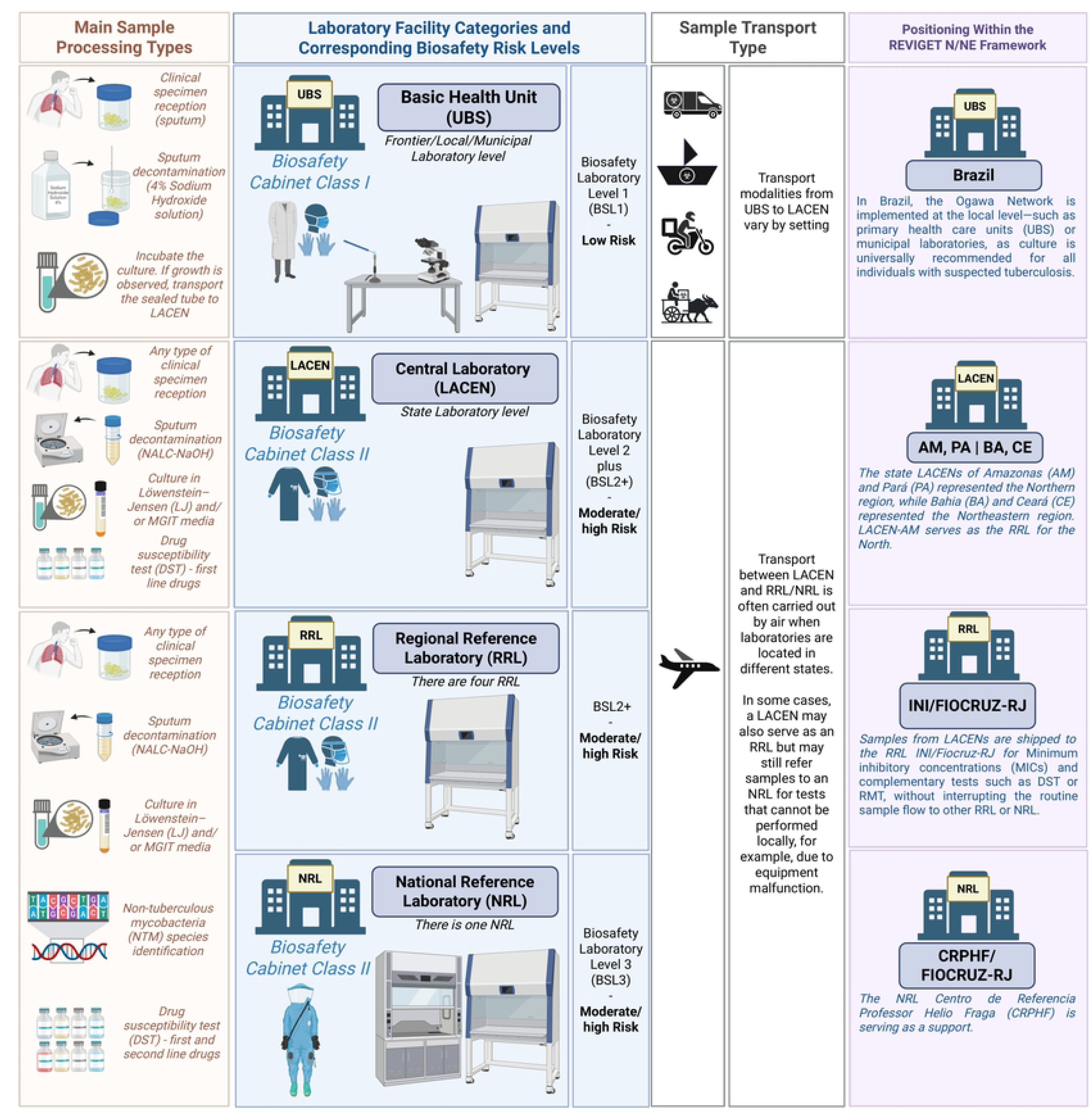
Schematic Representation of Sample Processing Complexity, Laboratory Biosafety Requirements, Transport Logistics, and Study Integration within the National Diagnostic and Surveillance Framework. Created in BioRender. Warren, R. (2025) https://BioRender.com/undefined.

In Brazil, a national program known as the Ogawa-Kudoh Network promotes universal culture for individuals with suspected tuberculosis. The Ogawa-Kudoh method, also referred to as the K-O swab method, is widely used for culturing *M. tuberculosis* from sputum samples, particularly in resource-limited settings. A critical step in this technique is sputum decontamination, which eliminates contaminating bacteria and debris while preserving viable mycobacteria. The procedure involves collecting sputum with a sterile swab, immersing it in a 4% sodium hydroxide solution for decontamination, and subsequently inoculating the swab onto Ogawa medium. Due to its operational simplicity, low cost, and minimal infrastructure requirements, this method is routinely implemented in laboratories at the local/municipal levels, such as the Basic Health Unit (UBS) (Brazil, 2022).

The disposal of biohazardous waste in this study will be according to Brazilian Law No. 12.305/2010, known as the National Solid Waste Policy (*Política Nacional de Resíduos Sólidos – PNRS*), Brazilian regulatory norm No. 12809 regarding health service waste manipulation and ANVISA regulatory norm (RDC) No. 222/2018, which specifically addresses the management of healthcare service waste. This legislation establishes guidelines for the integrated management and environmentally sound handling of solid waste, promoting practices such as waste reduction, reuse, recycling, and appropriate final disposal [27–29]. In addition, the National Health Surveillance Agency (ANVISA) and the National Council for the Environment (CONAMA) play key roles in overseeing and regulating laboratory waste disposal practices.

Culture transportation will follow the technical guidelines for biological risk, classifying the material as Risk Group B. It will be packaged and labelled with UN 2814 substance identification code, accompanied by the required documentation as specified in ANVISA’s Manual for Transport of Human Biological Material, in compliance with IATA regulations [30,31].

Administrative risks may include challenges in procurement, budgeting, and travel logistics. These will be addressed through collaboration with a dedicated and trained technical management team responsible for supporting project operations and ensuring smooth coordination across sites.

Risks to the institutional image could arise from procedural errors or infrastructural limitations that impact the execution of laboratory activities. These risks will be minimized through ongoing technical oversight and quality control. The project team has demonstrated experience in delivering high-quality training and has received strong performance evaluations in previous initiatives, which supports confidence in the implementation of this study.

Data management and privacy are also critical considerations. This study will follow the General Data Protection Law No. 13.709/2018 (*Lei Geral de Proteção de Dados Pessoais – LGPD*), which establishes regulations governing the collection, processing, storage, and sharing of personal data in Brazil. Its primary objective is to safeguard fundamental rights related to freedom, privacy, and the free development of the individual.

Mishandling of data could compromise patient confidentiality, and laboratory staff may face exposure to multidrug-resistant strains of *M. tuberculosis*. To mitigate these risks, all patient data will be anonymized, and access to sensitive information will be restricted to the study coordinator and designated team members using a secure REDCap database. This will be important for the identification of serial isolates and further investigation when necessary.

Lastly, the potential for misinterpretation of genomic and epidemiological correlations will be addressed through rigorous data analysis using appropriate statistical models and expert review. All interpretations will be collaboratively evaluated by the multidisciplinary research team to ensure scientific accuracy and alignment with the study objectives.

### 2.11 Project Status and Timeline

The project activities will be executed from 2024 to 2026. These activities are organized into ten Work Packages (WPs): WP1: Project Management and Data Infrastructure; WP2: Ethical and Epidemiological Requirements; WP3: Clinical Ecosystem; WP4: Laboratory Ecosystem; WP5: Bioinformatics and Digital Tools; WP6: Data Analysis; WP7: WGS Integration into RNDS and GEMIBRA; WP8: Development of Educational Materials; WP9: Capacity Building, Dissemination, and Scientific Communication; WP10: Political and Social Engagement. The timeline is detailed within the Supplementary Table 1.

### 2.12 Communication of Results, Health Promotion, and Community Engagement

Strategies for translating and disseminating knowledge about DR-TB diagnosis and treatment will be developed in partnership with civil society organizations, healthcare professionals, and health authorities. The project will promote health education and scientific outreach adapted to diverse audiences through educational activities, publications, and community engagement. Results will be disseminated through peer-reviewed journal articles, academic presentations, and public-facing events. These initiatives will take place concurrently with the laboratory and data management phases of the project, ensuring real-time communication and feedback.

## 3. Discussion

This study protocol outlines the implementation of a genomic epidemiological surveillance network (REVIGET) aimed at integrating WGS into routine TB diagnostics in Brazil, with a focus on DR-TB in the North and Northeast regions. These regions bear the highest DR-TB burden nationally yet face structural limitations in laboratory capacity, training, and timely diagnostics, resulting in suboptimal clinical outcomes and persistent transmission.

The practical application of genomic technologies within the Unified Health System (*Sistema Único de Saúde - SUS*) responds directly to strategic goals outlined in Brazil’s National Plan to End TB. By enabling local laboratories to perform WGS and interpret resistance profiles, the project supports the decentralization of high-complexity diagnostics and strengthens the capacity of state-level TB programs. These efforts align with global priorities for innovation, equity, and access to precision public health tools, particularly in resource-limited settings.

REVIGET’s implementation represents an important process innovation, incorporating WGS into DR-TB surveillance workflows. This has the potential to enhance case detection, accelerate treatment decisions, and support precision medicine by generating individualized resistance profiles and tracking transmission dynamics. Identifying novel mutations is highly important; however, understanding the mutations present in circulating strains is equally crucial, particularly for evaluating the sensitivity and specificity of new molecular methods in our setting. Moreover, integration with the National Health Data Network (RNDS) via the GEMIBRA platform will contribute to more timely, actionable insights across TB care continuously.

Scientific and technical outputs expected from this project include new insights into the genomic diversity and evolutionary pathways of MTBC, a better understanding of resistance mechanisms to newly introduced drugs (e.g., bedaquiline, delamanid, linezolid and pretomanid), and the generation of high-resolution epidemiological data in geographically historically underrepresented areas in TB genomics research. Our findings will be disseminated through peer-reviewed publications, technical manuals, scientific symposia and training events. The project also anticipates the formation of master’s and PhD students, contributing to long-term workforce development in genomics, surveillance, and bioinformatics.

Dissemination strategies will extend beyond scientific audiences to reach healthcare workers, patients, and the public. Materials such as printed guides, videos, and a mobile application (“TudoTB”) will promote health literacy and patient empowerment, particularly among vulnerable populations. Community engagement activities will involve civil society organizations, local TB committees, and public health authorities in participating states. These initiatives will address TB/HIV coinfection, treatment adherence, stigma situation, and healthcare access in historically underserved populations, including quilombola, riverside, Indigenous, and people deprived of liberty communities.

The project also promotes equity by ensuring representation and capacity-building in underserved regions, fostering inclusive research environments. Participatory approaches - such as local training, stakeholder meetings, and collaboration with advocacy committees - will guide the project’s implementation and help shape locally appropriate responses to DR-TB.

A potential limitation of this study is its reliance on culture-based workflows for WGS, which may lead to the exclusion of samples with delayed or failed growth, particularly in complex or extrapulmonary TB cases. This limitation could introduce bias in strain representation, potentially underestimating genomic diversity or specific resistance mutations. Moreover, implementation may face logistical constraints, including reagent shortages or delays in procurement and training across multiple regions. These risks will be addressed through contingency planning, centralized oversight and adaptive scheduling.

Preliminary results from the Brazilian Ministry of Health based on 2024 data demonstrated that between 2015 and 2024, a total of 20,628 new cases of DR-TB were reported in Brazil, including 1,047 cases registered in 2024. The historical trend reveals fluctuations over the years, with notable declines in 2016 and 2020, likely associated with stock shortages of the RMT Xpert MTB/RIF cartridges and the impact of the COVID-19 pandemic, respectively. From 2021 onward, a gradual recovery in case notifications was observed, peaking in 2022 with 1,231 reported cases, followed by a reduction in 2024 (n=1,047). The implementation of the RMT Network for TB in 2014 might have played a key role in this scenario, contributing significantly to the increased detection of RR-TB [32].

It is important to highlight that the spatial distribution analysis of newly diagnosed DR-TB cases between 2016 and 2024 indicates that all federal units (FUs) reported at least one case during this period. From 2016 to 2019, 3,875 new DR-TB cases were notified, with higher concentrations in the municipalities of Rio de Janeiro (n=547; 14.1%), Manaus (n=202; 5.2%), Porto Alegre (n=202; 5.2%), São Paulo (n=192; 5.0%), and Belém (n=156; 4.0%). These same cities also recorded the highest number of DR-TB cases from 2020 to 2024, accounting for a combined total of 5,170 new cases: Rio de Janeiro (n=651; 12.6%), Manaus (n=397; 7.7%), São Paulo (n=394; 7.6%), Belém (n=245; 4.7%), and Porto Alegre (n=176; 3.4%). Despite the negative impact of the COVID-19 pandemic on TB detection and follow-up, the number of DR-TB notifications increased by 33.4% from 2020 to 2024 compared to the period from 2016 to 2019 [32].

In 2023, the highest TB mortality rates in the country were recorded in the states of Amazonas (5.1 deaths per 100,000 population), Pernambuco (4.8 deaths per 100,000), and Rio de Janeiro (4.6 deaths per 100,000). Furthermore, 12 federal units reported mortality rates above the national average of 2.85 deaths per 100,000 population. The North region stood out with four states exhibiting mortality rates above 3.5 per 100,000, highlighting significant regional disparities in the response to the disease [32].

In 2024, TB incidence rates varied across the federal units as in previous years. The states with the highest incidence rates were Amazonas (94.7 cases per 100,000 population), Rio de Janeiro (75.3 cases per 100,000), and Roraima (64.3 cases per 100,000). Additionally, the North region concentrated five states with incidence rates above 50 cases per 100,000 population: Amazonas, Roraima, Pará (61.8), Acre (58.3), and Amapá (52.9), reinforcing the persistent regional inequalities in TB burden [32], which endorses initiatives as REVIGET N/NE, directed to attend the different regional aspects of TB dynamic within the complexity of a large and diverse country as Brazil.

Any protocol amendments, including modifications to sampling strategy, timelines, or analytical workflows, will be documented and submitted for ethical review through the Brazilian “*Plataforma Brasil”* system (https://plataformabrasil.saude.gov.br/login.jsf). If unforeseen challenges arise that compromise data integrity, participant safety, or the feasibility of project activities, study termination will be considered through consultation with institutional partners, funders, and ethics committees. Transparent reporting and documentation of any changes will be maintained throughout.

## Funding

This study was funded by the Brazilian National Council for Scientific and Technological Development (*Conselho Nacional de Desenvolvimento Científico e Tecnológico* - CNPq) grant process number 445784/2023-7 and by the Research Productivity Grants - PQ. CNPq Call No. 09/2022 Process 311165/2022-2. This project was also funded by Google Cloud (Project number: 209080321235).

## Competing interests

The authors have declared that no competing interests exist.

## Data availability

All data generated or analyzed during this study will be made publicly available upon completion of the study through the following repositories: GitHub (https://github.com/GEMIBRA and https://github.com/reviget), Zenodo (https://zenodo.org/communities/gemibra and https://zenodo.org/communities/reviget) and Open Science Framework (OSF) (https://osf.io/t562n/). Research outputs will be disseminated via Zenodo and OSF. It will include various materials such as datasets, peer-reviewed publications, conference abstracts, scientific protocols, educational resources, presentations, videos, books, and booklets. The retrospective dataset analyzed in this study is already publicly accessible through the National Center for Biotechnology Information (NCBI) under BioProject ID PRJNA1135144. All newly generated data from the current project will be deposited in the same repository and made publicly available following study completion and publication. Deidentified research data will be made accessible in an online repository upon publication. The study progress can be followed at the www.reviget.org website.

## Acknowledgements

The authors would like to express their sincere gratitude to the Brazilian Ministry of Health teams, General Coordination for Surveillance of Tuberculosis, Endemic Mycoses and Non-Tuberculous Mycobacteria (Coordenação-Geral de Vigilância da Tuberculose, Micoses Endêmicas e Micobactérias Não Tuberculosas - CGTM) and General Coordination of Public Health Laboratories (Coordenação-Geral de Laboratórios de Saúde Pública – CGLAB) for the invaluable technical and operational support to this study.

## Author’s Contributions

Conceptualization: ECC, KVBL and RMW. Methodology: ECC, KVBL, CCF, TCBB, AAF, DKCS, LEOF, AS, MCSL and RMW. Resources: KVBL, CCF, TCBB, AAF, AS and RMW. Writing - Original Draft: ECC and KVBL. Writing - Review & Editing: All authors. Visualization: ECC, RJPSG and RMW. Supervision: RMW and TO. Project administration: KVBL, ECC, CCF, TCBB and AAF. Funding acquisition: ECC, KVBL, CCF, TCBB, AAF, AS, DJM, LRR, RJPSG, IPF, RMW and TO. The REVIGET consortium members are described as follows. **Brazil, Pará team (26 members):** Alex Brito Souza^1,2^, Allana Aymê Farias Batalha^3^, Ana Judith Pires Garcia Quaresma^1^, Ana Paula Sousa Araújo^3^, Carlos Augusto Abreu Alberio^4^, Danna Karen Corrêa dos Santos^1,2^, Davi Josué Marcon^1,2^, Emilly Gabriele Ribeiro Dias^1,5^, Emmily Oliveira Amador^1,2^, Erilene Cristina da Silva Furtado^3^, Gleissy Adriane Lima Borges^3^, Ismari Perini Furlaneto^6^, José Anderson Monteiro Pantoja^1^, Juan Pereira Soares^1^, Karla Valéria Batista Lima^1,2,5^, Katia Cristina de Lima Furtado^3^, Layana Rufino Ribeiro^1,5^, Lúcia Helena Martins Tavares Monteiro^7^, Luna Luana de Jesus Pantoja^3^, Maria Isabel Montoril Gouveia^1,5^, Patricia Miriam Sayuri Sato Barros da Costa^3^, Rafaella Bonfim Barros^3^, Rosa Márcia Saraiva Gentil^3^, Ricardo José de Paula Souza e Guimarães^8^, Valnete das Graças Dantas Andrade^3^, Victor Cezar Gomes Melo^3^**. Brazil, Amazonas team (13 members):** Adriana Ayden Ferreira^9^, Claudio Fernández Araujo^9^, Etelvina das Graças da Costa Zaranza^9^, Gustavo Kiyoshi Massunari^9^, Katiuscia Dantas de Araújo^9^, Layssa do Carmo Barroso^9^, Lilian Eduarda de Oliveira Freitas^9^, Marco Aurélio Almeida de Oliveira^9^, Marlucia da Silva Garrido^9^, Reinaldina Dorotheia Nascimento Vieira^9^, Renata Lia Coragem Carvalho^9^, Tatyana Costa Amorim Ramos^9^, Walter André Junior^9^. **Brazil, Bahia team (11 members):** Andrezza Kariny Miranda de Souza^10^, Erivelton Oliveira^11^, Felicidade Mota Pereira^11^, Gabriela Sant’Ana Menezes de Andrade^11^, Hendor Neves Ribeiro de Jesus^10^, Ingrid Pimentel Silva Gabriel^10^, Marcela Kelly Astete Gomez^11^, Rodrigo Alves da Costa Goncalves^10^, Theolis Costa Barbosa Bessa^10, 12^, Tonya Azevedo Duarte^10^, Vanessa Brandão Nardy^11^. **Brazil, Ceará team (13 members):** Alex Pessoa da Rocha^13^, Ana Karoliny Alves^14^, Antonia Cely Vitor Barbosa^13^, Cristiane Cunha Frota^14^, Karene Ferreira Cavalcante^13^, Larissa Leão Ferrer de Sousa^13^, Michelle Guilherme de Lima^13^, Rosiane Marcelino Lobo Fernandes^13^, Shirlene Telmos Silva de Lima^13^, Sidarta Lopes Viana^13^, Soraya de Oliveira Sancho^13^, Thiago Pinto Brasil^14^, Vânia Angelica Feitosa Viana^13^. **Brazil, Rio de Janeiro team (6 members):** Leonardo Bruno Paz Ferreira Barreto^15, 16^, Maria Cristina da Silva Lourenço^16^, Yrneh Prado Palacios^16^, Philip Noel Suffys^17^, Fátima Fandinho^18^, William Marco Vicente da Silva^18^. **South Africa, Stellenbosch University team (4 members):** Abhinav Sharma^19^, Emilyn Costa Conceição^12,19,20^, Robin Mark Warren^19^, Tulio de Oliveira^20^. The consortium members’ affiliations are as follows.

1. Secao de Bacteriologia, Instituto Evandro Chagas, Ananindeua, Pará, Brazil.

2. Programa de Pos-Graduação em Biologia Parasitaria na Amazonia, Universidade do Estado do Para, Belem, Brazil.

3. Laboratorio Central do Estado do Para, Belem, Para, Brazil.

4. Hospital Universitario Joao de Barros Barreto, Ambulatorio de Tuberculose Multiressistente, Universidade Federal do Para, Belem, Para, Brazil.

5. Programa de Pos-graduação em Epidemiologia e Vigilancia em Saude, Instituto Evandro Chagas, Ananindeua, Brazil.

6. Centro Universitario do Para, Belem, Para, Brazil.

7. Secretaria de Estado da Saude do Para, Belem, Para, Brazil.

8. Secao de Geoprocessamento, Instituto Evandro Chagas, Ananindeua, Para, Brazil.

9. Laboratorio Central do Amazonas, Fundação de Vigilancia em Saude do Amazonas Dra Rosemary Costa Pinto, Manaus, Amazonas, Brazil.

10. Fundação Oswaldo Cruz, Instituto Gonçalo Moniz, Salvador, Bahia, Brasil.

11. Laboratorio Central do Estado da Bahia, Salvador, Bahia, Brazil.

12. Rede Brasileira de Pesquisas em Tuberculose, Rio de Janeiro, Rio Janeiro, Brazil.

13. Laboratorio Central do Estado do Ceara, Fortaleza, Ceara, Brazil.

14. Departamento de Patologia e Medicina Legal, Faculdade de Medicina, Universidade Federal do Ceara, Fortaleza, Ceara, Brazil.

15. Laboratorio de Bacteriologia e Bioensaios em Micobacterias, Instituto Nacional de Infectologia Evandro Chagas, Fundacao Oswaldo Cruz, Manguinhos, Rio de Janeiro, Rio de Janeiro, Brazil.

16. Programa de Pos-Graduacao em Pesquisa Clinica e Doencas Infecciosas, Instituto Nacional de Infectologia Evandro Chagas, Fundacao Oswaldo Cruz, Manguinhos, Rio de Janeiro, Rio de Janeiro, Brazil.

17. Laboratorio de Biologia Molecular Aplicada a Micobacterias, Instituto Oswaldo Cruz, Fundacao Oswaldo Cruz, Rio de Janeiro, Brazil.

18. Laboratorio de Referencia Nacional de Tuberculose e Micobacterioses Angela Maria Werneck, Centro de Referencia Professor Helio Fraga, Escola Nacional de Saude Publica, Fundaçao Oswaldo Cruz, Rio de Janeiro, Brazil.

19. South African Medical Research Council Centre for Tuberculosis Research, Division of Molecular Biology and Human Genetics, Faculty of Medicine and Health Sciences, Stellenbosch University, Cape Town, South Africa.

20. Centre for Epidemic Response and Innovation, School of Data Science and Computational Thinking, Stellenbosch University, Stellenbosch, South Africa.

## Supporting Information

Supplementary Table 1. Phases of the Study and Planned Timeline of Activities Over 24 Months.

## Notes

### Competing Interest Statement

The authors have declared no competing interest.

### Funding Statement

Yes

### Author Declarations

This study was submitted for ethical evaluation through the Brazilian system (Plataforma Brasil) and addressed to the Research Ethics Committee (CEP) of the led institution, IEC, which was approved No 7,282,078 and Certificate of Presentation of Ethical Appreciation (CAAE): 85134924.4.0000.0019. This project was also registered per Brazilian Law No. 13,123/2015 and its regulations, through the National System for the Management of Genetic Heritage and Associated Traditional Knowledge (SisGen), under Registration No. A247AD9.

## References

1. Transforming our world: the 2030 Agenda for Sustainable Development | Department of Economic and Social Affairs. [cited 2 May 2025]. Available: https://sdgs.un.org/2030agenda

2. World Health Organization. The end TB strategy. World Health Organization, editor. Global Tuberculosis Programme; 2015. Available: https://iris.who.int/bitstream/handle/10665/331326/WHO-HTM-TB-2015.19-eng.pdf?sequence=1

3. Huang C-T, Tsai Y-J, Shu C-C, Lei Y-C, Wang J-Y, Yu C-J, et al. Clinical significance of isolation of nontuberculous mycobacteria in pulmonary tuberculosis patients. Respir Med. 2009;103: 1484–1491. doi:10.1016/j.rmed.2009.04.017

4. Ojo O, Odeyemi A. Non-Mycobacteria Tuberculosis in Africa: A Literature Review. Ethiop J Health Sci. 2023;33: 913–918. doi:10.4314/ejhs.v33i5.21

5. Ying C, Zhang L, Jin X, Zhu D, Wu W. Advances in diagnosis and treatment of non-tuberculous mycobacterial lung disease. Diagn Microbiol Infect Dis. 2024;109: 116254. doi:10.1016/j.diagmicrobio.2024.116254

6. Sibandze DB, Kay A, Dreyer V, Sikhondze W, Dlamini Q, DiNardo A, et al. Rapid molecular diagnostics of tuberculosis resistance by targeted stool sequencing. Genome Med. 2022;14: 52. doi:10.1186/s13073-022-01054-6

7. Pham J, Su LD, Hanson KE, Hogan CA. Sequence-based diagnostics and precision medicine in bacterial and viral infections: from bench to bedside. Curr Opin Infect Dis. 2023;36: 228–234. doi:10.1097/QCO.0000000000000936

8. Maljkovic Berry I, Melendrez MC, Bishop-Lilly KA, Rutvisuttinunt W, Pollett S, Talundzic E, et al. Next Generation Sequencing and Bioinformatics Methodologies for Infectious Disease Research and Public Health: Approaches, Applications, and Considerations for Development of Laboratory Capacity. J Infect Dis. 2020;221: S292–S307. doi:10.1093/infdis/jiz286

9. Zhang H, Liu M, Fan W, Sun S, Fan X. The impact of Mycobacterium tuberculosis complex in the environment on one health approach. Front Public Health. 2022;10: 994745. doi:10.3389/fpubh.2022.994745

10. Vogel M, Utpatel C, Corbett C, Kohl TA, Iskakova A, Ahmedov S, et al. Implementation of whole genome sequencing for tuberculosis diagnostics in a low-middle income, high MDR-TB burden country. Sci Rep. 2021;11: 15333. doi:10.1038/s41598-021-94297-z

11. WHO. Catalogue of mutations in Mycobacterium tuberculosis complex and their association with drug resistance. 1st ed. Geneva: World Health Organization - WHO; 2022. Available: https://www.who.int/publications-detail-redirect/9789240028173

12. WHO. Catalogue of mutations in Mycobacterium tuberculosis complex and their association with drug resistance, second edition. Geneva: World Health Organization - WHO; 2023.

13. WHO. Global Tuberculosis Report 2024. World Health Organization - WHO; 2024. Available: https://www.who.int/teams/global-tuberculosis-programme/tb-reports/global-tuberculosis-report-2024

14. Goldenberg T, Gayoso R, Mogami R, Lourenço MC, Ramos JP, Carvalho LD de, et al. Clinical and epidemiological characteristics of M. kansasii pulmonary infections from Rio de Janeiro, Brazil, between 2006 and 2016. J Bras Pneumol Publicacao Of Soc Bras Pneumol E Tisilogia. 2020;46: e20190345. doi:10.36416/1806-3756/e20190345

15. Lima AC de O de, Schmid KB, Melo HF de, Athayde RC, Monte RL, Almeida IN de, et al. Molecular characterization of nontuberculous Mycobacteria in a tuberculosis and HIV reference unit in the State of Amazonas, Brazil. Rev Soc Bras Med Trop. 2022;55: e0613. doi:10.1590/0037-8682-0613-2021

16. Barboza GCS, Almeida IN de, Santos LBD, Augusto CJ, Leal ÉA, Pádua CAM de, et al. Nontuberculous mycobacteria in patients of a specialty hospital. Rev Inst Med Trop Sao Paulo. 2023;65: e42. doi:10.1590/S1678-9946202365042

17. Sousa E de O, Carneiro RT de O, Montes FCOF, Conceição EC, Bartholomay P, Marinho JM, et al. Laboratory-based study of drug resistance and genotypic profile of multidrug-resistant tuberculosis isolates in Salvador, Bahia, Brazil. Rev Soc Bras Med Trop. 2022;55: e00132022. doi:10.1590/0037-8682-0013-2022

18. Conceição EC, Salvato RS, Gomes KM, Guimarães AE dos S, da Conceição ML, Souza e Guimarães RJ de P, et al. Molecular epidemiology of Mycobacterium tuberculosis in Brazil before the whole genome sequencing era: a literature review. Mem Inst Oswaldo Cruz. 2021;116. doi:10.1590/0074-02760200517

19. Meehan CJ, Goig GA, Kohl TA, Verboven L, Dippenaar A, Ezewudo M, et al. Whole genome sequencing of Mycobacterium tuberculosis: current standards and open issues. Nat Rev Microbiol. 2019;17: 533–545. doi:10.1038/s41579-019-0214-5

20. Mesquita CR, Conceição EC, Monteiro LHMT, da Silva OM, Lima LNGC, de Oliveira RAC, et al. A Clinical-Epidemiological and Geospatial Study of Tuberculosis in a Neglected Area in the Amazonian Region Highlights the Urgent Need for Control Measures. Int J Environ Res Public Health. 2021;18: 1335. doi:10.3390/ijerph18031335

21. CLSI M24: SUSCEPTIBILITY TESTING. CLSI M24: susceptibility testing of mycobacteria, nocardia spp., and other aerobic actinomycetes. Place of publication not identified: GOBI US LIBRARY SOLUTIONS; 2018.

22. Conceição EC, Wells F, Williams J, Ismail N, john.metcalfe, Rie A van, et al. Mycobacterium tuberculosis DNA Extraction Using InstaGene Matrix and High Speed Homogenizer from Clinical P… 2024 [cited 18 May 2025]. Available: https://www.protocols.io/view/mycobacterium-tuberculosis-dna-extraction-using-in-dhvs366e

23. Heupink TH, Verboven L, Sharma A, Rennie V, Fuertes M de D, Warren RM, et al. The MAGMA pipeline for comprehensive genomic analyses of clinical Mycobacterium tuberculosis samples. PLOS Comput Biol. 2023;19: e1011648. doi:10.1371/journal.pcbi.1011648

24. Brasil. NR 32 - Segurança e Saúde no Trabalho em Serviços de Saúde. Ministério do Trabalho e Emprego; 2005. Available: https://www.gov.br/trabalho-e-emprego/pt-br/assuntos/seguranca-e-saude-no-trabalho/normatizacao/normas-regulamentadoras/norma-regulamentadora-n-32-nr-32

25. Brasil. Diretrizes Gerais para o Trabalho em Contenção com Material Biológico. Ministério da saúde, Secretaria de Ciência, Tecnologia e Insumos estratégicos; 2004. Available: https://bvsms.saude.gov.br/bvs/publicacoes/diretrizes_trabalho_material_biologico.pdf

26. Brasil. Manual de Recomendações para o Diagnóstico Laboratorial de Tuberculose e Micobactérias não Tuberculosas de Interesse em Saúde Pública no Brasil. 1st ed. Brasília: Ministério da Saúde; 2022.

27. Brasil. Lei n° 12.305, de 2 de agosto de 2010. Presidência da República; 2010. Available: http://www.planalto.gov.br/ccivil_03/_ato2007-2010/2010/lei/l12305.htm

28. ABNT. NBR 12809:2013 – Gestão de resíduos de serviços de saúde – Gerenciamento intraestabelecimento. Associação Brasileira de Normas Técnicas; 2013.

29. ANVISA. RDC n° 222, de 28 de março de 2018. Agência Nacional de Vigilância Sanitária (ANVISA); 2018.

30. Brasil. Riscos Biológicos: Guia Técnico. Ministério do Trabalho e Emprego; 2008.

31. ANVISA. Manual para o Transporte de Material Biológico Humano. Agência Nacional de Vigilância Sanitária; 2008. Available: https://www.gov.br/anvisa/pt-br/assuntos/publicacoes/publicacoes/vigilancia-sanitaria/manual-de-transporte-de-material-biologico-humano

32. Brasil. Boletim Epidemiológico - Tuberculose 2025. Ministério da Saúde, Secretaria de Vigilância em Saúde, Departamento de Doenças de Condições Crônicas e Infecções Sexualmente Transmissíveis, Coordenação-Geral de Vigilância das Doenças de Transmissão Respiratória de Condições Crônicas; 2025. Available: https://www.gov.br/aids/pt-br/central-de-conteudo/boletins-epidemiologicos/2025/boletim-epidemiologico-tuberculose-2025/view

